# Missense variants in *TUBA4A* cause myo-tubulinopathies

**DOI:** 10.1101/2025.06.26.25330266

**Authors:** Mridul Johari, Chiara Folland, Yoshihiko Saito, Machteld M. Oud, Jevin M. Parmar, Ana Töpf, Sergei Kurbatov, Maria Ampleeva, Ekaterina Y. Zakharova, Irina A. Chekmareva, Ksenia S. Shirokova, Dmitrii Atiakshin, Thatjana Gardeitchik, Erik-Jan Kamsteeg, Evita Medici, Laura Donker Kaat, Christine C. Bruels, Seth A. Stafki, Elicia A. Estrella, Hannah R. Littel, Louis M. Kunkel, Peter B. Kang, Ikeoluwa Osei-Owusu, Lynn Pais, Melaine O’Leary, Christina Austin-Tse, Anne O’Donnell-Luria, Brian Mangilog, Francesca Clementina Radio, Adele D’Amico, Andrea Ciolfi, Marco Tartaglia, Aurélien Perrin, Charles Van Goethem, Guilhem Sole, Marie-Laure Martin-Négrier, Mireille Cossée, Casie A. Genetti, Zaheer M. Valivullah, Vedrana Milic, Gordana Kovacevic, Ana Kosac, Cristiane A.M. Moreno, Clara Gontijo Camelo, Edmar Zanoteli, Michael C. Fahey, Alan H. Beggs, John Vissing, Volker Straub, Marco Savarese, Giorgio Tasca, Nicol Voermans, Nigel G. Laing, Bjarne Udd, Ichizo Nishino, Gianina Ravenscroft

## Abstract

Tubulinopathies encompass a wide spectrum of disorders resulting from variants in genes encoding α- and β-tubulins, the key components of microtubules. While previous studies have linked *de novo* or dominantly inherited *TUBA4A* missense variants to neurodegenerative phenotypes, including amyotrophic lateral sclerosis, frontotemporal dementia, hereditary spastic ataxia, and more recently, an isolated report of congenital myopathy, the full phenotypic and genotypic spectrum of *TUBA4A*-related disorders remains incompletely characterised. In this multi-centre study, we identified 13 novel *TUBA4A* missense variants in 31 individuals from 19 unrelated families. Remarkably, affected individuals in 17 families presented with a primary axial myopathy without any identified CNS involvement or history of such disease. In the remaining two families, we observed probands with cerebellar ataxia and epilepsy accompanying proximal and axial muscle weakness, establishing the first documented association between *TUBA4A* variants and multisystem proteinopathy.

Our cohort exhibited diverse genotypes and associated inheritance patterns: four families demonstrated autosomal dominant transmission through heterozygous variants in *TUBA4A*, three probands had homozygous *TUBA4A* variants, where the biallelic genotype was found to be associated with the disease, and the heterozygous carriers were asymptomatic; five probands carried *de novo* variants, and nine probands with heterozygous *TUBA4A* variants were classified as "isolated-sporadic cases" where parental samples were unavailable. Clinical phenotypes ranged from mild to severe myopathy, predominantly affecting the axial and paraspinal muscles. We observed a range of disease onset, from congenital to late adulthood. Creatine kinase levels were also variable, ranging from normal to highly elevated. Cardiac function remained preserved across the cohort.

Muscle biopsies revealed a range of pathologies, including myofibre size variation, myofibre atrophy, nemaline bodies, core-like regions, internal nuclei, and endomysial fibrosis. Immunohistochemical staining showed evidence of proteinopathy, with autophagic features and TUBA4A accumulation in patient myofibres. Complementary *in silico* and *in vitro* investigations suggested that the identified *TUBA4A* substitutions cause significant protein abnormalities and may differentially impact microtubule dynamics.

Our findings establish myo-tubulinopathies as distinct clinical entities, encompassing both primary myopathies and multisystem proteinopathies with muscle involvement. This study broadens the phenotypic and genotypic spectrum of *TUBA4A*-related disorders beyond autosomal dominant or *de novo* mechanisms and neurodegenerative presentations. These results underscore the importance of considering *TUBA4A* variants in the differential diagnosis of axial myopathies and multisystem proteinopathies, regardless of central nervous system (CNS) involvement.

## Introduction

Tubulinopathies arise from defective microtubule function or assembly due to pathogenic variants in the genes encoding different tubulin isotypes.^1^ The term ‘tubulinopathy’ was first used to describe diseases associated with brain malformations arising from defects in cortical development and faulty neuronal migration, which are at the end of the phenotypic spectrum.^1^ However, ‘tubulinopathies’ are now an umbrella term for a wide spectrum of ciliary, zygotic arrest and early developmental failure, neurological and neurodegenerative phenotypes caused by pathogenic defects in microtubule units and associated proteins.

Microtubules, a fundamental cytoskeletal component in all forms of life, exhibit dynamic instability—a continuous cycle of growth and shrinkage that is essential for various cellular functions, including maintaining cell shape, enabling intracellular transport, and participating in cell division.^2–4^ Microtubules are composed of different tubulin subunits encoded by several genes, resulting in various tubulin isotypes. The most common of these, α- and β-tubulin, form a noncovalent heterodimer, which serves as the building blocks of microtubules.^5,6^ In humans, nine genes encode different α-tubulin isotypes, each of which can potentially pair with any of the ten β-tubulin isotypes, contributing to microtubule diversity and functional specialisation. Amongst the nine α-tubulin isotypes, *TUBA4A* encodes tubulin alpha 4a. Smith and colleagues first observed *TUBA4A*-related microtubule disarray in patients with familial amyotrophic lateral sclerosis (ALS), demonstrating that *TUBA4A* mutants have an impaired ability to form α- and β-tubulin dimers, which disrupts microtubule dynamics and stability through a dominant-negative mechanism.^7^ Later, pathogenic variants in *TUBA4A* were reported in patients with frontotemporal dementia (FTD),^8,9^ Parkinson’s disease,^10^ hereditary spastic ataxia,^11,12^ and zygotic arrest.^13,14^ Recently, a study identified a *de novo* variant in *TUBA4A* associated with an isolated congenital myopathy phenotype in two unrelated probands.^15^

The pathogenic variants observed so far in *TUBA4A* are not restricted to any domain; how these variants result in a specific phenotype remains unknown. Impaired dimerisation and microtubule instability are shared functional defects of most mutant tubulin proteins, including *TUBA4A* mutants.^7,16–19^ However, these gain-of-function pathomechanisms, primarily due to missense variants, can lead to vastly different phenotypes.^7,12^ Additionally, heterozygous protein truncating variants in *TUBA4A*, which escape nonsense-mediated decay and result in loss of parts of C-terminal TUBA4A, ultimately forming a shorter protein, have also been associated with ALS and FTD phenotypes.^7,20^

In our study, we present a gamut of *TUBA4A*-related primary myopathies and multisystem proteinopathies, providing detailed clinical and functional insights. We identified 13 variants in 31 affected individuals from 19 unrelated families. We observed autosomal dominant, including *de novo*, and recessive inheritance. Disease onset ranged from congenital to late adulthood, predominantly affecting axial and proximal muscles. Notably, unlike other tubulinopathies, 17 out of 19 families neither exhibited any central nervous system impairment nor had a history of such disease. In the remaining two cases, multisystem involvement of myopathy, cerebellar ataxia and epilepsy were seen. Through *in silico*, *in vitro*, histopathological and immunohistochemical analyses, we characterise the phenotypic and molecular spectrum of *TUBA4A*-related myopathies and multisystem proteinopathies – henceforth referred to as ‘myo-tubulinopathies’.

## Materials and Methods

### Patients and clinical examinations

All probands and affected members underwent clinical neuromuscular and neurological examinations. The inclusion criteria for this cohort were a presumptive diagnosis of myopathy and the presence of a rare coding variant in *TUBA4A,* as identified through diagnostic or research testing. We collected blood and/or saliva samples for DNA extraction from affected and unaffected members from 19 families. Muscle imaging was obtained as part of the routine diagnostic work-up and was available for inclusion in this study for some patients, with consent. Electrophysiological examination results, including nerve conduction studies and needle electromyography (EMG), as well as serum creatine kinase (CK) measurements, were obtained for most patients. Aggregated clinical data was available for 21 patients. The studies were conducted per the Declaration of Helsinki, and ethical approval was obtained through institutional review boards (Supplementary Data 1(available upon request)).

### Muscle biopsy and immunohistochemical studies

Muscle biopsies were obtained from a subset of affected patients (*n*=13). Routine muscle histopathological studies were performed, including hematoxylin and eosin (H&E) staining, modified Gomori trichrome staining, and NADH tetrazolium reductase (NADH-TR) staining.^21^ DAB immunostaining was performed using mouse monoclonal anti-myotilin (clone RSO34, 1:20, LEICA Biosystems Newcastle Ltd, UK) and mouse monoclonal anti-desmin (clone D33, 1:70, Richard-Allan Scientific, USA), with mouse ExtrAvidin Peroxidase Staining Kit (EXTRA2, Merck KGaA, Darmstadt, Germany). Microscopic images were obtained using a NIKON ECLIPSE Ci microscope equipped with an OLYMPUS ColorView II camera. Additional staining for aggregate pathology was performed on patient samples (Patients 5, 8, 9, 11, 12, and 24, *n = 6*). TUBA4A immunostaining was performed using a rabbit monoclonal anti-TUBA4A antibody (clone EPR13477(B), 1:100, Abcam, UK) after heat-mediated antigen retrieval with citrate buffer (pH 6.0) for 8 min at 91°C. SQSTM1 (p62) immunostaining used mouse monoclonal anti-SQSTM1 antibody (clone D-3, 1:200, Santa Cruz Biotechnology, Inc, USA). TARDBP (TDP-43) staining was performed using a rabbit polyclonal anti-TDP-43 antibody (1:1,000, Proteintech, Japan) after heat-mediated antigen retrieval with TE buffer (pH 9.0) for 8 minutes at 95°C.

### Electron microscopy and quantification analysis

Ultrathin resin sections (70-80 nm) were prepared for electron microscopy from patients 8, 9, 25 and 26 and two healthy control individuals and examined using a FEI Tecnai Spirit Transmission Electron Microscope operating at 120 kV. Electron micrographs were obtained using the Gatan Digital Micrograph (Gatan, Inc, USA).

For microtubule quantification, ultrathin resin sections from Pat.8 and Pat.9 were used. Ten microtubules were identified in these two patients and only five in controls. Average diameters were calculated, and an unpaired t-test was used for statistical calculations.

### DNA sequencing and analysis

Targeted gene panel sequencing, short read (sr)-exome sequencing (ES) and sr-genome sequencing (GS) were carried out on probands and available family members (Supplementary Data 1(available upon request)). Sanger sequencing was performed to confirm the variants identified in the probands and co-segregate the variants with disease in available family members.

Single nucleotide variant (SNV) and indel analyses were done through a) seqr- a web-based platform hosted by the Centre for Population Genomics (CPG; Garvan Institute of Medical Research, Murdoch Children’s Research Institute) and the Centre for Mendelian Genomics (CMG; the Broad Institute),^22^ b) RD-Connect Genome Phenome Analysis Platform (GPAP),^23^ and custom in-house pipelines using a minor allele frequency (MAF) ≤ 0.0001 in the Genome Aggregation Database (gnomAD) v2.1.1 (hg19),^24^ and v3.1.2 (hg38).^25^

### *TUBA4A* variant analysis and interpretation

Variants in *TUBA4A* were annotated on NM_006000.3 (ENST00000248437.9, MANE Select) and NP_005991.1 (protein). Variant frequency reanalysis was performed using the updated gnomAD v4.1.0.

Identified variants were assessed using various *in silico* tools (Supplementary Data 2) per current ACMG/AMP guidelines. The variant assessment was refined using the germline variant classifier VarSome,^26^ with a modified pathogenicity interpretation based on the evidence levels available, including those collected during this study.

### *In-silico* modelling of *TUBA4A* variants

#### Multiple sequence alignment

To examine the conservation of substituted amino acid positions, we performed multiple sequence alignments of TUBA4A orthologs across different species and human TUBA4A paralogs. TUBA4A ortholog and paralog protein sequences were retrieved from UniProt using their respective accession codes. Alignments were performed using the default MAFFT algorithm in Jalview (v2.11.2).

#### Calculation of free energy changes and TUBA4A aggregation prediction

We used FoldX (v5.0)^27^ to investigate the impact of *TUBA4A* variants on protein stability and to show the free energy changes between the wild-type and mutant proteins. Free energy change predictions were only performed on missense substitutions within high confidence regions (pLDDT > 90) of the AlphaFold2-predicted protein structure.^28^ The AlphaFold2-derived wild-type TUBA4A protein structure, based on NM_006000.3, was energy-minimised and prepared for mutagenesis with the ‘RepairPDB’ and ‘BuildModel’ commands in FoldX, respectively. Subsequent free energy calculations were performed with the ‘Stability’ command, and differences in free energy were determined by subtracting the free energy value of the wild-type protein monomer from that of the mutant protein (ΔG_MUT_ - ΔG_WT_). Calculations were replicated 10 times with default parameters for each substitution. Substitutions were classified as having a destabilising effect on protein folding if the free energy difference (ΔΔG) was >1.6 kcal/mol.^27,29,30^ Furthermore, we calculated the significance of FoldX stability predictions per-variant by comparing the per-variant values against a conservative destabilisation threshold of 1.6 kcal/mol using a bootstrapping approach. Analyses and plotting were performed in RStudio (v2024.12.1+563) R (v4.4.3).

Aggrescan 4D was used to examine the aggregation propensity of wild-type and mutant TUBA4A.^31,32^ Since there is no TUBA4A 3D protein structure derived from X-ray diffraction or solution NMR, Aggrescan utilises modelling approaches in PDB/mmCIF format to predict the aggregation propensities of TUBA4A. We used the A4D Dynamic mode, which integrates the CABS-flex tool to model protein flexibility and incorporate this information into the assessment of aggregation properties.^33^ In this approach, the simulation begins with a FoldX- optimised input structure as the starting point.^27^ Aggregation propensity scores were calculated for all 448 amino acids of TUBA4A across both wild-type and mutant proteins. The normalised average aggregation propensity score of each TUBA4A mutant protein was calculated relative to wild-type, using the Aggrescan4D-derived scores. Analyses and plotting were performed in RStudio (v2023.06.1) R (v4.3.3).

#### TUBA4A structural modelling and *in silico* mutagenesis

The predicted wild-type TUBA4A protein structure, based on NM_006000.3/NP_005991.1, was retrieved from the AlphaFold Protein Structure Database^34^ using its UniProt accession code (P68366). Three-dimensional protein structures were visualised using PyMol (v. 2.5.2). To examine the effects of variants on three-dimensional protein structure, *in silico* mutagenesis for identified missense substitutions was performed through the PyMol ‘mutagenesis’ function. To examine whether the variants affect any binding sites, an experimentally validated model of mouse TUBA4A-TUBB2A with kinesin (PDB: 8IXF)^35^ was visualised, and images were exported using Mol*.^36^

### *In vitro* over-expression of *TUBA4A* variants in COS1 cells

Wild-type human *TUBA4A* ORF was cloned into pcDNA3 to produce an N-terminally HA-tagged construct. The observed missense variants were introduced to the construct by site-directed mutagenesis. Wild-type and mutant *TUBA4A* were overexpressed in COS1 cells to study the localisation of TUBA4A and microtubule morphology. COS1 cells were cultured in Dulbecco’s Modified Eagle Medium (DMEM) supplemented with 10% fetal calf serum (FCS). Cells were grown on glass coverslips and, at approximately 60% confluence, transfected with Lipofectamine 2000 (Invitrogen) according to the manufacturer’s procedure. Cells were transfected with either WT or *TUBA4A*-myopathy variants: p.(Thr179Ile), p.(Leu227Phe), p.(Ser241Phe), p.(Glu254Ala), p.(Glu254Gln), p.(Glu254Lys), p.(Glu254Gly), p.(Gly354Asp). Additionally, we also transfected cells with previously identified TUBA4A ALS-causing substitution p.(Arg105Cys) and p.(Arg320Cys).^7^ Upon 48 hours of incubation, the cells were fixed with ice-cold methanol for 10 minutes at room temperature (RT) and blocked with 2% bovine serum albumin (BSA) in PBS, followed by 1-hour incubation with the primary anti-HA antibody (mouse monoclonal, Sigma-Aldrich, Zwijndrecht, Netherlands), a PBS wash and another 1 hr incubation with the secondary anti-mouse Alexa Fluor-488 antibody (ThermoFisher Scientific Waltham, USA). The coverslips were embedded in Fluoromount-D with DAPI (Southern Biotech, Birmingham, AL, USA) on a microscopic glass slide. Then, the coverslips were analysed at 63x magnification with a Zeiss Axio Imager Z2 microscope (Zeiss, Sliedrecht, Netherlands) equipped with an ApoTome slider. The experiments were performed in triplicate. The statistical difference for the abnormal pattern was calculated for the wildtype compared to the TUBA4A-mutants using a two-way ANOVA followed by Tukey’s post hoc test.

## Data availability statement

All relevant imaging and biological materials included in this study are available from the authors (or commercial providers). Sensitive sr-ES/sr-GS data of patients and family members are deposited in seqr and RD-Connect GPAP. Due to the extensive clinical details in Supplementary Data 1, which may contain potentially identifying information, this section has been removed from the preprint version. Readers interested in the detailed case-level data should contact the corresponding author for access, subject to appropriate data sharing agreements and ethical guidelines.

## Results

### Clinical features in myo-tubulinopathies

A summary of clinical findings from 21 affected individuals is provided in Table 1. Detailed clinical descriptions are provided in the (Supplementary Data 1(available upon request)). In this myo-tubulinopathy cohort, the mean age of onset was 15.1 years (range: 0 – 60 years). Four patients had onset of neuromuscular symptoms at birth. Two patients died during the first year of life (Pat.9 and Pat.10). Within the cohort, 56% (*n* = 10/18) were non-ambulant. Most patients exhibited mild to moderate proximal and distal muscle weakness. Proximal upper and lower limb strength was more frequently affected than distal strength. While more than half of the cohort had normal to mildly reduced distal limb strength, a smaller subset showed moderate to severe weakness, particularly in the lower limbs (Table 1).

**Table 1:** Summary of clinical features of affected individuals in our myo-tubulinopathy cohort with disease causing variants in TUBA4A. Each feature is shown as x/y under "Total", where ’x’ is the number of individuals exhibiting the feature, and y is the number of individuals for whom data were available in that category. The corresponding percentage is calculated from these values. Denominators vary due to missing data.

|  | Individuals with disease causing variants in TUBA4A (Number of patients, n=21, Number of families, N=19) |  |
| --- | --- | --- |
|  | Total | Percentage |
| **Inheritance (families)** |  |  |
| AD | 4/19 | 21% |
| AR | 3/19 | 16% |
| de novo | 5/19 | 26% |
| Sporadic* | 7/19 | 47% |
| *inheritance for Pat. 14 is suspected paternal but cannot be determined. |  |  |
| **Onset** |  |  |
| At birth | 4/21 | 19% |
| Childhood | 11/21 | 52% |
| Adult and late adult (18-30) | 2/21 | 10% |
| Mid or late onset (30-60) | 4/21 | 19% |
| **Biological Sex** |  |  |
| Male | 11/21 | 52% |
| Female | 10/21 | 48% |
| **Muscle weakness (MRC 4-5, 2-3, 0-1)** |  |  |
| Distal UL |  |  |
| MRC 4-5 | 11/20 | 55% |
| MRC 2-3 | 4/20 | 20% |
| MRC 0-1 | 5/20 | 25% |
| Distal LL |  |  |
| MRC 4-5 | 11/20 | 55% |
| MRC 2-3 | 4/20 | 20% |
| MRC 0-1 | 5/20 | 25% |
| Proximal UL |  |  |
| MRC 4-5 | 7/20 | 35% |
| MRC 2-3 | 8/20 | 40% |
| MRC 0-1 | 5/20 | 25% |
| Proximal LL |  |  |
| MRC 4-5 | 7/20 | 35% |
| MRC 2-3 | 7/20 | 35% |
| MRC 0-1 | 6/20 | 30% |
| Assymetry | 2/21 | 10% |
| Non-ambulant* | 10/18 | 56% |
| *two patients died in first year of life |  |  |
| Axial weakness | 18/19 | 95% |
| Facial weakness | 9/20 | 40% |
| Ptosis | 6/20 | 25% |
| Respiratory involvement | 15/19 | 79% |
| **Histopathology** |  |  |
| Fiber size variation | 13/13 | 100% |
| Internal nuclei | 11/13 | 85% |
| Autophagic vacuoles | 4/13 | 31% |
| Myofibrillar disorganisation | 4/13 | 31% |
| p62 positive accumulations | 5/5 | 100% |
| TDP43 positive accumulations | 3/3 | 100% |
| Nemaline bodies | 4/13 | 31% |

A representation of muscle strength and weakness in our cohort, based on the MRC scale, is shown in Fig. 1A. An asymmetrical muscle weakness pattern was observed in 9% of patients. Axial muscle weakness was a consistent feature in our myo-tubulinopathy cohort (18/19). Nearly half of the patients also showed facial muscle involvement, and a smaller proportion (25%) presented with ptosis. Notably, ophthalmoplegia was identified in two individuals, and ophthalmoparesis in one — an uncommon finding in myopathies. Respiratory muscle weakness was observed in most cases (79%). Serum CK data was available for 21 individuals in the cohort, and three of these patients had normal CK. The mean level was 1,468.5 IU/l (range: 143-2,794 IU/l). For patients where muscle MRIs were available (Fig. 1B), fatty degeneration of gluteus minimus and erector spinae and often preserved gluteus maximus muscles. Accordingly, axial and paraspinal muscle weakness was observed as the most common feature.

**Fig. 1:**
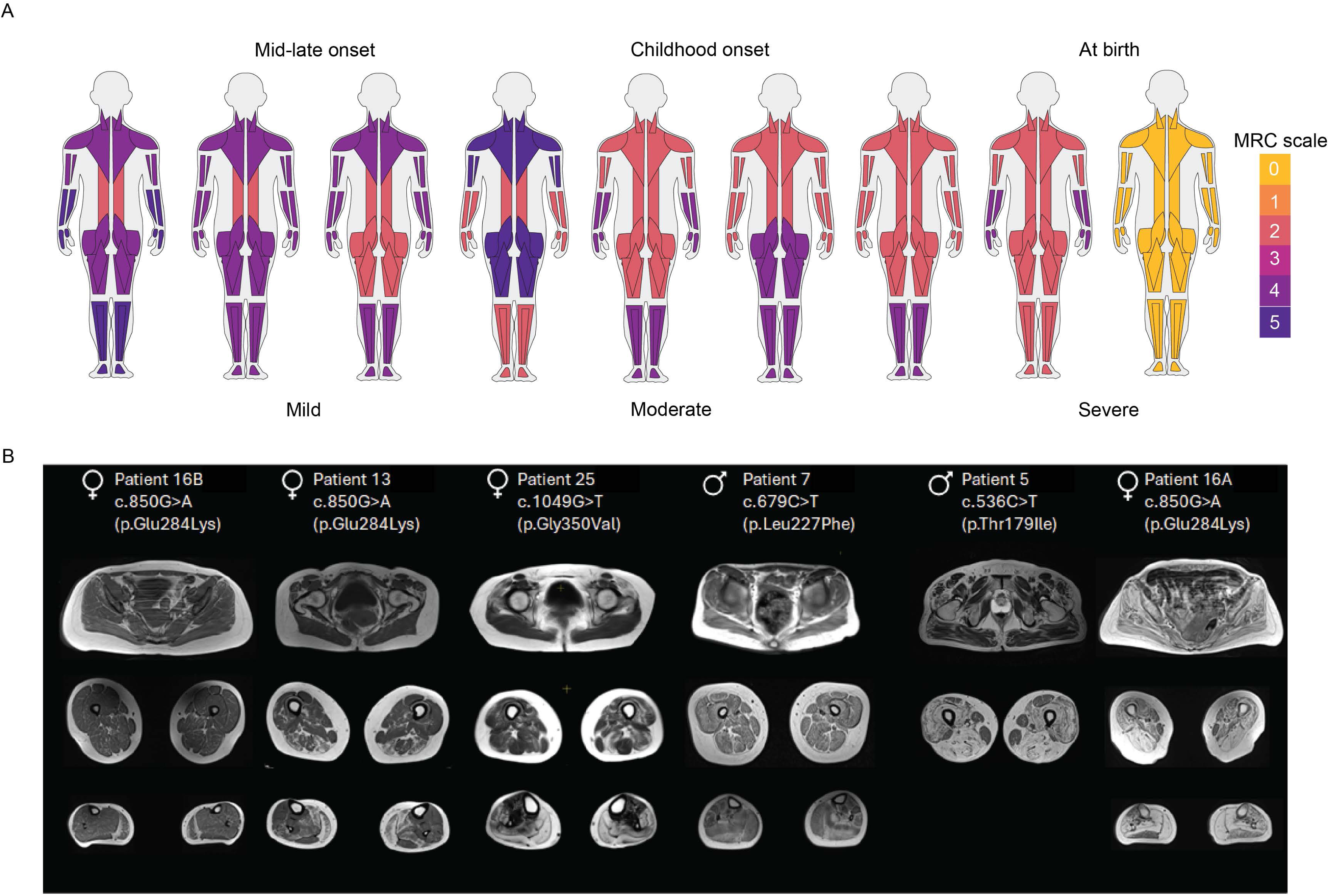
*TUBA4A*-related myopathy presents with variable patterns of muscle weakness, most consistently affecting axial muscles. A) Spectrum of muscle weakness observed in patients with variants in *TUBA4A*. MuscleViz (https://muscleviz.github.io) was used to visualise the weakness using the MRC scale for muscle strength. Muscle weakness patterns and age of onset observed in our myo-tubulinopathy cohort are shown by the body diagrams. B) Axial T1-weighted MR images of muscles of the gluteal region, thighs, and calves from six patients with *TUBA4A* variants, arranged from left to right in order of increasing disease severity. Each panel includes the patient’s sex, age at imaging, and specific *TUBA4A* variant.

In 17 out of 19 families, at the last assessment, no clinical signs or history of a neurodegenerative disease including hereditary spastic ataxia, ALS, dementia or Alzheimer disease were observed or stated. However, in two patients we observed multisystem involvement presenting as cerebellar ataxia and/or epileptic seizures along with a myofibrillar myopathy with prominent axial muscle weakness.

### Muscle pathology and protein accumulation pattern in myo-tubulinopathies

Histopathological findings, where detailed data was available (*n* = 13), are summarised separately in Table 2, all muscle biopsies showed marked variation in myofibre size. Otherwise, there were no unifying histopathological hallmarks across the cohort. Eleven biopsies exhibited internally located nuclei, six showed predominance of type I myofibres and nine revealed endomysial fibrosis (Fig. 2A). Additionally, autophagic vacuoles and myofibrillar disorganisation were observed in four biopsies each.

**Fig. 2:**
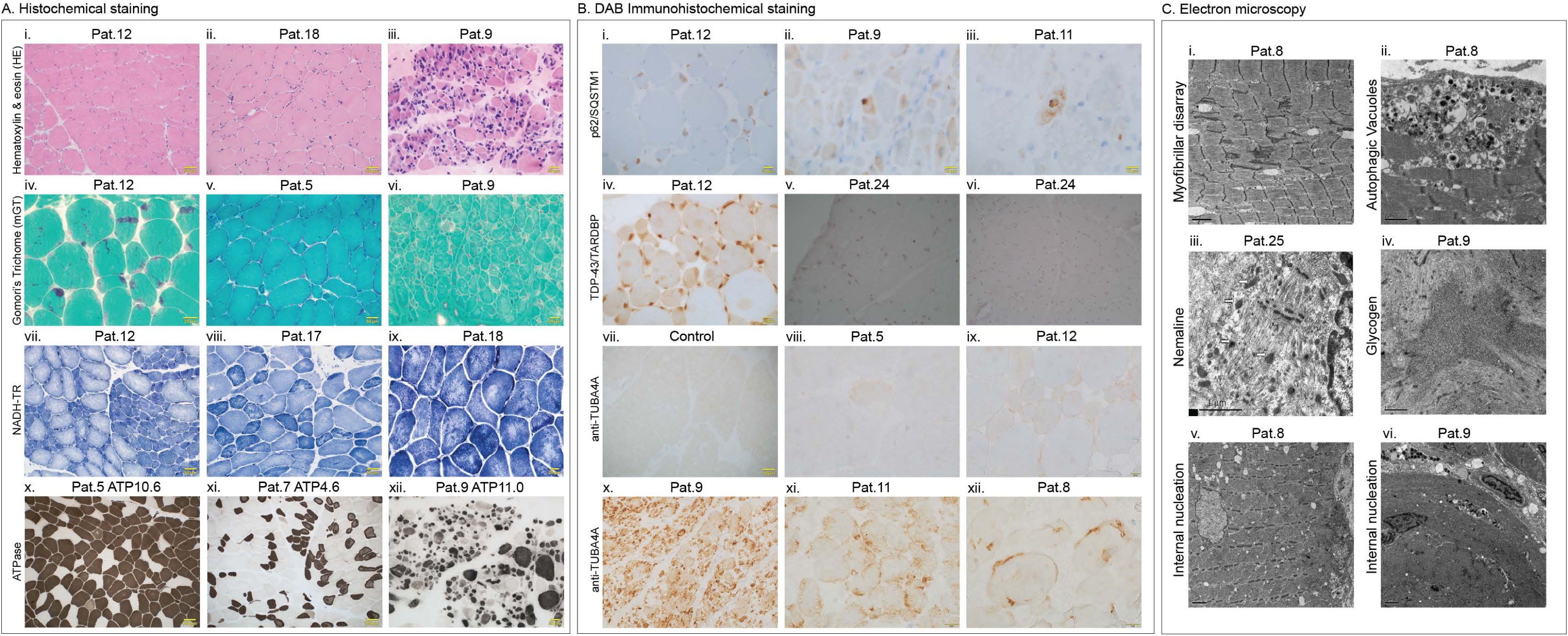
*TUBA4A*-related myopathy shows a spectrum of histological, immunohistochemical and ultrastructural findings. A) (i-ix) Histochemical staining of patient muscle biopsies: i-iii) Hematoxylin & eosin (H&E) stain showing myofibre size variation, small angular myofibres, internal nucleation and endomysial fibrosis. iv-vi) Modified Gomori’s trichome staining showing presence of Nemaline bodies, spheroid or cytoplasmic bodies and occasional rimmed vacuoles. vii-ix) NADH-TR staining showing myofibre type and marked atrophy of type 1 myofibres along with minicores, large cores and moth eaten myofibres. x-xii) ATPase staining showing variability in myofibre size and atrophy, type 1 myofibre predominance and immature myofibres. B. (i-ix) DAB Immunohistochemical staining of patient and control muscle biopsies: i-iii) Increased immunoreactivity for p62/SQQSTM1 staining in all cases examined. iv-vi) However, TDP-43/TARDBP staining showed variable immunoreactivity for available muscle samples. vii-xii) Staining for anti-TUBA4A also showed variable immunoreactivity in different patients as compared to control muscle sample. C. (i-vi) Electron microscopy of patient muscles: i-ii) Ultrastructural analysis of Pat.8 muscle biopsy showing myofibrillar disarray and autophagic vacuoles. iii-iv) Pat.25 showed presence of Nemaline bodies while glycogen accumulation is seen in Pat.9. v-vi) Internal nucleation is seen in muscle biopsies of both Pat.8 and Pat.9.

**Table 2:** Histopathological features observed in individuals from our myo-tubulinopathy cohort.

|  | *Variation in Fiber size* | *Internal nucleation* | *Type I Fiber predominance* | *Endomysial Fibrosis* | *Autophagic Vacuoles* | *Nemaline bodies* | *Spheroid bodies* | *Cores* | *Myofibrillar disorganisation* |
| --- | --- | --- | --- | --- | --- | --- | --- | --- | --- |
| Pat.5 | Y | Y | N | Y | N | N | Y | Y | N |
| Pat.8 | Y | Y | N | Y | Y | N | N | N | Y |
| Pat.9 | Y | Y | Y | Y | Y | Y | Y | N | N |
| Pat.11 | Y | Y | N | Y | N | N | N | N | N |
| Pat.12 | Y | Y | Y | Y | N | Y | N | N | N |
| Pat.13 | Y | N | Y | N | N | N | N | N | N |
| Pat.15 | Y | Y | N | Y | N | N | N | Y | Y |
| Pat.16 | Y | Y | N | N | N | N | N | N | N |
| Pat.17 | Y | Y | N | Y | N | N | Y | N | N |
| Pat.18 | Y | Y | Y | Y | N | N | Y | N | N |
| Pat.24 | Y | N | N | N | Y | N | N | Y | Y |
| Pat.25 | Y | Y | N | Y | N | Y | Y | N | N |
| Pat.26 | Y | Y | Y | N | Y | Y | N | N | Y |

To further investigate accompanying proteinopathy features, immunohistochemical analysis was performed in a subset of patients. All examined cases (*n*=4) demonstrated increased immunoreactivity for p62/SQSTM1 staining, indicating enhanced accumulation of this protein in myofibres. In contrast, TDP-43/TARDBP staining showed pronounced phosphorylated TDP-43 accumulations in some biopsies and not in others. Similarly, anti-TUBA4A staining revealed heterogenous levels of TUBA4A positive accumulations in patients’ biopsies when compared to control muscle samples, which may reflect differences in the underlying variants (Fig. 2B).

Additionally, a subset of patients also exhibited peculiar ultrastructural features. Nemaline bodies were identified in four patients (Pat.9, Pat.12, Pat.25 and Pat.26, Fig. 2A-iv, Fig. 2C-iii). Other rare features included spheroid bodies, multiminicores, moth-eaten myofibres, abnormal glycogen accumulation, and small angular myofibres (Fig. 2A, 2C).

### Identification of variants in *TUBA4A*

Within the cohort we identified 13 different missense *TUBA4A* variants in 19 families. A representation of the observed variants in *TUBA4A* is depicted in Fig. 3A. Autosomal dominant (*n*=4 families), *de novo* (*n*=5 probands) and autosomal recessive (*n*=3 probands) variants were observed. In nine sporadic cases with heterozygous variants the mode of inheritance could not be clarified due to lack of additional familial samples. At least in one family (Pat.14), the deceased parent of the proband was indicated as affected. Aggregated genetic findings are summarised in Table 3. Interestingly, in four unrelated cases, four different heterozygous variants were identified which impacted the same amino-acid residue, Glu254, resulting in substitutions p.(Glu254Gly) and p.(Glu25Lys), confirmed to have arisen *de novo* in the probands, and p.(Glu254Ala) and p.(Glu254Gln) in two sporadic cases. Two of these variants, p.(Glu254Gln) and p.(Glu254Gly) were identified in the patients who died within the first year of life.

**Fig. 3:**
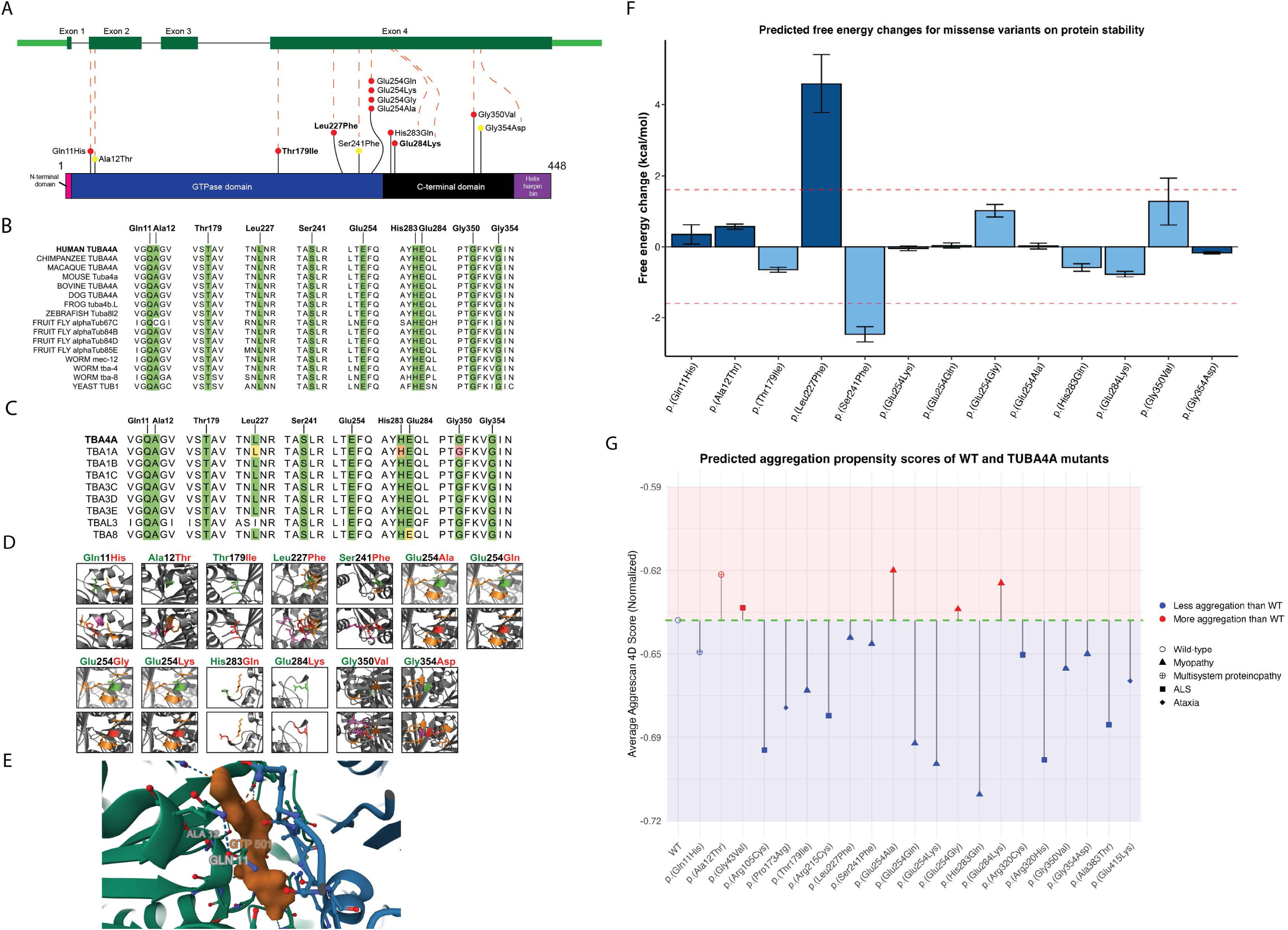
Modelling of variants in *TUBA4A* and *in silico* assessment of their impact suggests distinct pathomechanisms associated with different *TUBA4A* variants. A) Gene model of *TUBA4A* and 2D model of TUBA4A protein, showing the position of different myopathy variants (indicated as ‘coloured lollipops’) observed in our cohort. p.(Thr179Ile), p.(Leu227Phe) and p.(Glu284Lys) are in bold to indicate being observed in multiple families. p.(Ala12Thr), p.(Ser241Phe) and p.(Gly354Asp) are indicated with a ‘yellow lollipop’ to indicate homozygosity. Domain information for TUBA4A protein is obtained from Interpro-CATH gene3D. Exon 1 codes for the initiation site – methionine which forms the N-terminal domain. Thereafter, 2-268 amino acids form the GTPase domain, 269- 383 form the C-terminal domain and 384-448 form the Helix hairpin bin. All our observed variants are in the GTPase domain and the C-terminal domain. B) Conservation of Tubulins across species showed that all identified variants in our myopathy cohort are at highly conserved sites. C) Conservation of identified TUBA4A variant amino acid residues across other human tubulin paralogs. Conserved amino acids are highlighted with a green box. Pathogenic variants (red box), likely pathogenic variants (orange box), and variants of uncertain significance (yellow box) identified in paralogous human tubulins are also shown. D) The effect of identified missense variants on three-dimensional TUBA4A structure. Wild-type residues are coloured green and mutant residues are coloured red. Residues with which the mutant variant shows steric hindrance are purple. Residues which have polar contacts with the wild-type or variant residues are orange. E) A snapshot of mouse TUBA4A-TUBB2A with kinesin (PDB: 8IXF) using Mol*. The GTP molecule (orange) is shown along with TUBA4A (green) and TUBB2A (blue). Gln11 and Ala12 residues are labelled close to the tubulin catalytic GTP site. Gln11 is shown interacting with the GTP molecule. F) Predicted free energy changes (ΔG_MUT_ – ΔG_WT_) for missense variants on protein stability. Calculations were performed for variants within very high confidence regions (pLDDT > 90, dark blue bars) or confident regions (70 < pLDDT < 90, light blue bars) of the AlphaFold2-generated TUBA4A protein. Data are given as mean ± s.d. The red dashed lines indicate conservative thresholds (±1.6 kcal/mol). G) Aggrescan 4D prediction plot recreated with Aggrescan 4D scores showing aggregation propensity of wild type TUBA4A and TUBA4A variants including 13 variants identified in the present study for myopathy and multisystem proteinopathy, six variants associated with ALS and two variants previously published in association with ataxia. The x-axis represents the residue position, and the y-axis represents the normalised average aggregation propensity scores calculated by Aggrescan 4D.

**Table 3:**
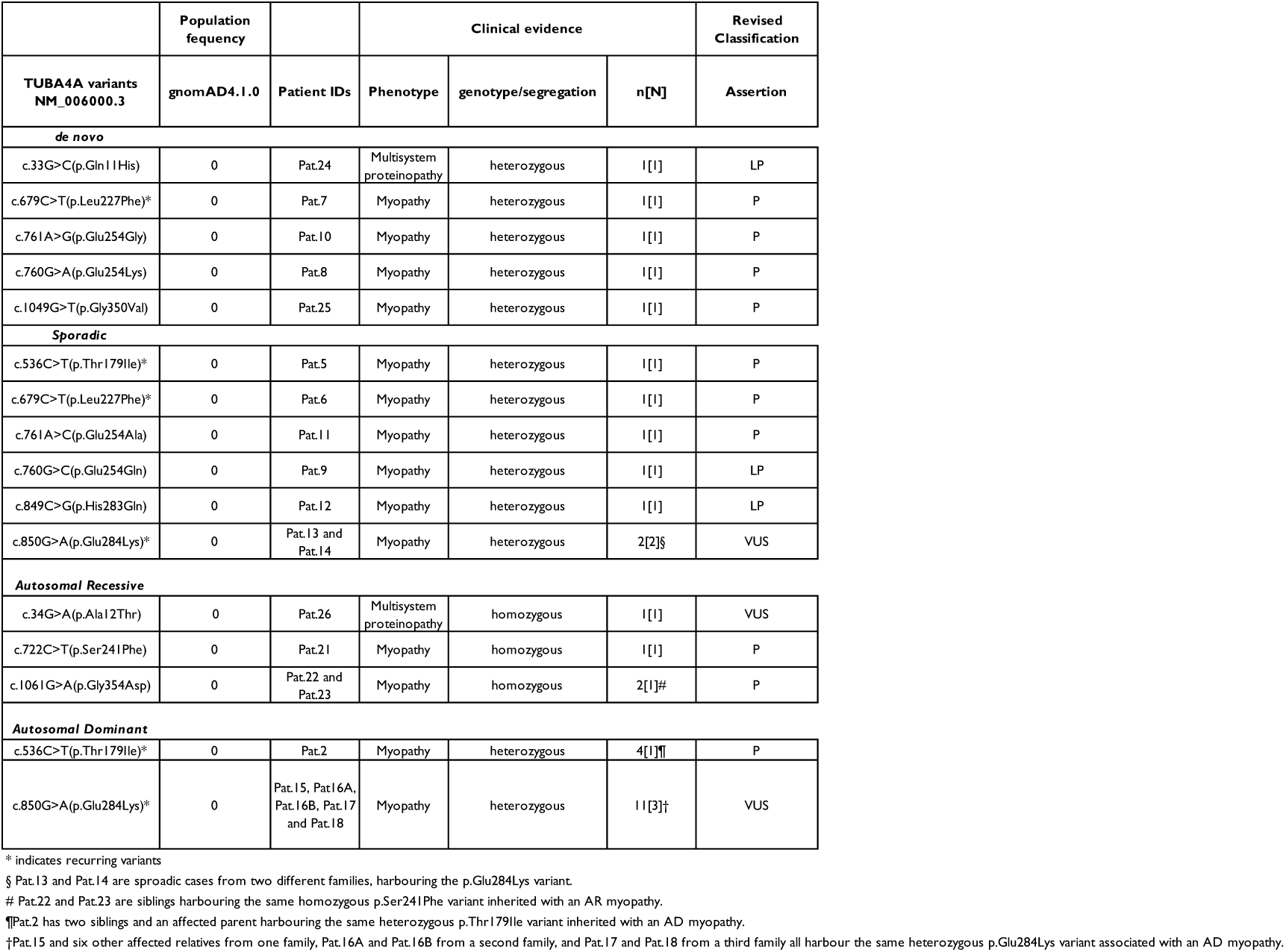
Assessment of the thirteen TUBA4A variants observed in our myo-tubulinopathy cohort. AR = Autosomal recessive, AD = Autosomal dominant, n=number of confirmed affected individuals (31), N=number of families (19).

|  | Population frequency |  | Clinical evidence |  |  | Revised Classification |
| --- | --- | --- | --- | --- | --- | --- |
| TUBA4A variants NM_006000.3 | gnomAD4.1.0 | Patient IDs | Phenotype | genotype/segregation | n[N] | Assertion |
| **de novo** |  |  |  |  |  |  |
| c.33G>C(p.Gln11His) | 0 | Pat.24 | Multisystem proteinopathy | heterozygous | 1[1] | LP |
| c.679C>T(p.Leu227Phe)* | 0 | Pat.7 | Myopathy | heterozygous | 1[1] | P |
| c.761A>G(p.Glu254Gly) | 0 | Pat.10 | Myopathy | heterozygous | 1[1] | P |
| c.760G>A(p.Glu254Lys) | 0 | Pat.8 | Myopathy | heterozygous | 1[1] | P |
| c.1049G>T(p.Gly350Val) | 0 | Pat.25 | Myopathy | heterozygous | 1[1] | P |
| **Sporadic** |  |  |  |  |  |  |
| c.536C>T(p.Thr179Ile)* | 0 | Pat.5 | Myopathy | heterozygous | 1[1] | P |
| c.679C>T(p.Leu227Phe)* | 0 | Pat.6 | Myopathy | heterozygous | 1[1] | P |
| c.761A>C(p.Glu254Ala) | 0 | Pat.11 | Myopathy | heterozygous | 1[1] | P |
| c.760G>C(p.Glu254Gln) | 0 | Pat.9 | Myopathy | heterozygous | 1[1] | LP |
| c.849C>G(p.His283Gln) | 0 | Pat.12 | Myopathy | heterozygous | 1[1] | LP |
| c.850G>A(p.Glu284Lys)* | 0 | Pat.13 and Pat.14 | Myopathy | heterozygous | 2[2]§ | VUS |
| **Autosomal Recessive** |  |  |  |  |  |  |
| c.34G>A(p.Ala12Thr) | 0 | Pat.26 | Multisystem proteinopathy | homozygous | 1[1] | VUS |
| c.722C>T(p.Ser241Phe) | 0 | Pat.21 | Myopathy | homozygous | 1[1] | P |
| c.1061G>A(p.Gly354Asp) | 0 | Pat.22 and Pat.23 | Myopathy | homozygous | 2[1]# | P |
| **Autosomal Dominant** |  |  |  |  |  |  |
| c.536C>T(p.Thr179Ile)* | 0 | Pat.2 | Myopathy | heterozygous | 4[1]¶ | P |
| c.850G>A(p.Glu284Lys)* | 0 | Pat.15, Pat.16A, Pat.16B, Pat.17 and Pat.18 | Myopathy | heterozygous | 11[3]† | VUS |

\* indicates recurring variants
§ Pat.13 and Pat.14 are sporadic cases from two different families, harbouring the p.Glu284Lys variant.

One heterozygous variant, p.(Glu284Lys), was identified in three unrelated families and two sporadic cases, with a total of 13 affected individuals harbouring this variant. In two probands, the same substitution, p.(Leu227Phe), was observed, one confirmed to be *de novo* and the other in a sporadic case. This same substitution was recently identified to have arisen *de novo* in two myopathy probands in China.^15^

Three *TUBA4A* variants were observed in homozygosity in our myo-tubulinopathy cohort. Firstly, the two families, Pat.21 and the siblings Pat.22 and Pat.23 with the homozygous variants, p.(Ser241Phe) and p.(Gly354Asp), respectively, had disease onset in the first year of life but this was not fatal in the first year. The parents of these three patients were heterozygous carriers and did not manifest any symptoms of neuromuscular disease. In family of Pat. 26, the proband, with an age of onset at 56-60 years, was observed to be homozygous for variant p.(Ala12Thr). His child was found to be heterozygous carrier of the variant and at last clinical assessment had idiopathic epilepsy without any neuromuscular issues. A healthy sibling of the proband had a normal genotype (Supplementary Data 1(available upon request)).

All identified variants were in exon 4, except for p.(Gln11His) and p.(Ala12Thr) which were within exon 2. Nine variants were in the guanosine triphosphate (GTP)-ase domain, and the remaining four variants were in the C-terminal domain (Fig. 3A). All 13 variants were absent in population databases including gnomADv4.1.0 (Supplementary Data 2).

### Structural modelling and aggregation analyses reveal variant-specific molecular impacts on TUBA4A

Multiple sequence alignments were performed for TUBA4A orthologs across 10 non-human species (Fig. 3B), and all human alpha tubulins (Fig. 3C), anchored to the human TUBA4A protein sequence. All the identified myo-tubulinopathy variants affected highly conserved amino acid residues.

The wild-type TUBA4A protein was predicted by AlphaFold2. The ‘mutagenesis’ function on PyMol was used to predict changes in protein structure and/or folding upon incorporating substituted amino acid residues into the sequence (Fig. 3D). Most substitutions showed a change in steric hindrance with nearby or distant amino acid residues. Of note, the two variants observed as *de novo*, p.(Leu227Phe), p.(Gly350Val) and the homozygous variant p.(Gly354Asp) showed the most substantial changes to polar contacts and steric hindrance upon substitution. Thus, we predicted that these substitutions could affect protein stability. Free energy change analyses showed that, of the identified substitutions, only the p.(Gln11His), p.(Ala12Thr), p.(Leu227Phe), and p.(Gly354Asp) were in the very high confidence regions (pLDDT > 90) of the AlphaFold2-predicted protein structure. FoldX-calculated free energy changes for these substitutions indicated that only the p.(Leu227Phe) substitution has a significant decrease in free energy > 1.6 kcal/mol, suggesting a protein destabilising effect with high-confidence (Fig. 3F). This aligns with the previously observed steric hindrance caused by p.(Leu227Phe) (Fig. 3D).

In addition, the p.(Ser241Phe) substitution showed a significant increase in free energy. This may predict a stability increase, but it cannot be excluded that the variants could cause a deleterious effect to protein folding or stability by another unknown mechanism.

Additionally, we used the experimentally validated model of mouse TUBA4A-TUBB2A dimer with kinesin (PDB: 8IXF) to visualise variant impact on the GTP binding site. In the α/β-tubulin heterodimer, both subunits can bind GTP; however, only the β-tubulin subunit is capable of hydrolysing GTP to GDP, while the GTP bound to α-tubulin (TUBA4A) remains non-exchangeable and structurally stabilising. The Gln11 and Ala12 residues in TUBA4A are in proximity to the non-exchangeable GTP-binding site, with Gln11 directly interacting with the nucleotide (Fig. 3E). We predict that substitutions at these positions, which alter the residue charge or polarity, may impair GTP binding and thereby destabilise the TUBA4A structure or its interaction with the β-tubulin, thus affecting the microtubule dynamics, an underlying pathomechanisms of different tubulinopathies.

Given that structural destabilisation alone may not fully capture the pathogenic potential of the identified variants, we subsequently investigated the aggregation propensity of TUBA4A variants, as stability and aggregation are distinct yet interconnected properties. While FoldX evaluates thermodynamic stability of the native protein fold, aggregation analysis specifically examines the potential for proteins to form multi-protein assemblies with potentially non-native conformations.

A variant may significantly destabilise the structure (high ΔΔG in FoldX) without promoting aggregation if it lacks aggregation-prone motifs or is managed by the chaperones.^37,38^ Conversely, even stable variants can expose or create aggregation-prone regions, altering cellular behaviour.^39^ By evaluating both factors, we aimed to clarify how TUBA4A substitutions might contribute to the pathobiology through distinct molecular mechanisms. Aggrescan 4D predicts whether individual residues are aggregation prone (positive score, > 0), have increased solubility (negative score, < 0) or no change in solubility is predicted (0) (Supplementary Data 2). The dynamic mode also predicts the change in interacting residues and the aggregation propensity of the whole protein. Thus, the average aggregation score represents the average of all 448 residues of TUBA4A in both wild-type and each variant protein. All tested variants, including the 13 identified variant residues in our study and the eight previously observed ALS/ataxia-causing variants alter the aggregation propensity of the protein in a variable manner (Fig. 3G, Supplementary Data 2).

These findings suggest that distinct variants, despite differing in predicted stability and aggregation propensity, may modulate TUBA4A behaviour through diverse mechanisms.

### *In-vitro* studies show variant-specific disruption of TUBA4A localisation and microtubule morphology

To experimentally evaluate the effect of the missense substitutions on intracellular localisation of TUBA4A, COS1 cells were transfected with either WT or TUBA4A variants: p.(Arg105Cys), p.(Thr179Ile), p.(Leu227Phe), p.(Ser241Phe), p.(Glu254Ala), p.(Glu254Gln), p.(Glu254Lys), p.(Glu254Gly), p.(Arg320Cys) and p.(Gly354Asp). Overexpression of wild-type TUBA4A led to an abnormal TUBA4A localisation pattern in ∼53% of the cells. A similar result was seen for p.(Glu254Gly) (∼47%) and p.(Glu254Lys) (∼56%), while p.(Glu254Ala) and p.(Glu254Gln) showed less abnormal cells, ∼23% and ∼26% respectively. A near complete abnormal localisation pattern was observed in ∼87-100% of the cells (Fig. 4A) over-expressing p.(Thr179Ile), p.(Leu227Phe), p.(Ser241Phe), p.(Arg320Cys) and p.(Gly354Asp) localisation.

**Fig. 4:**
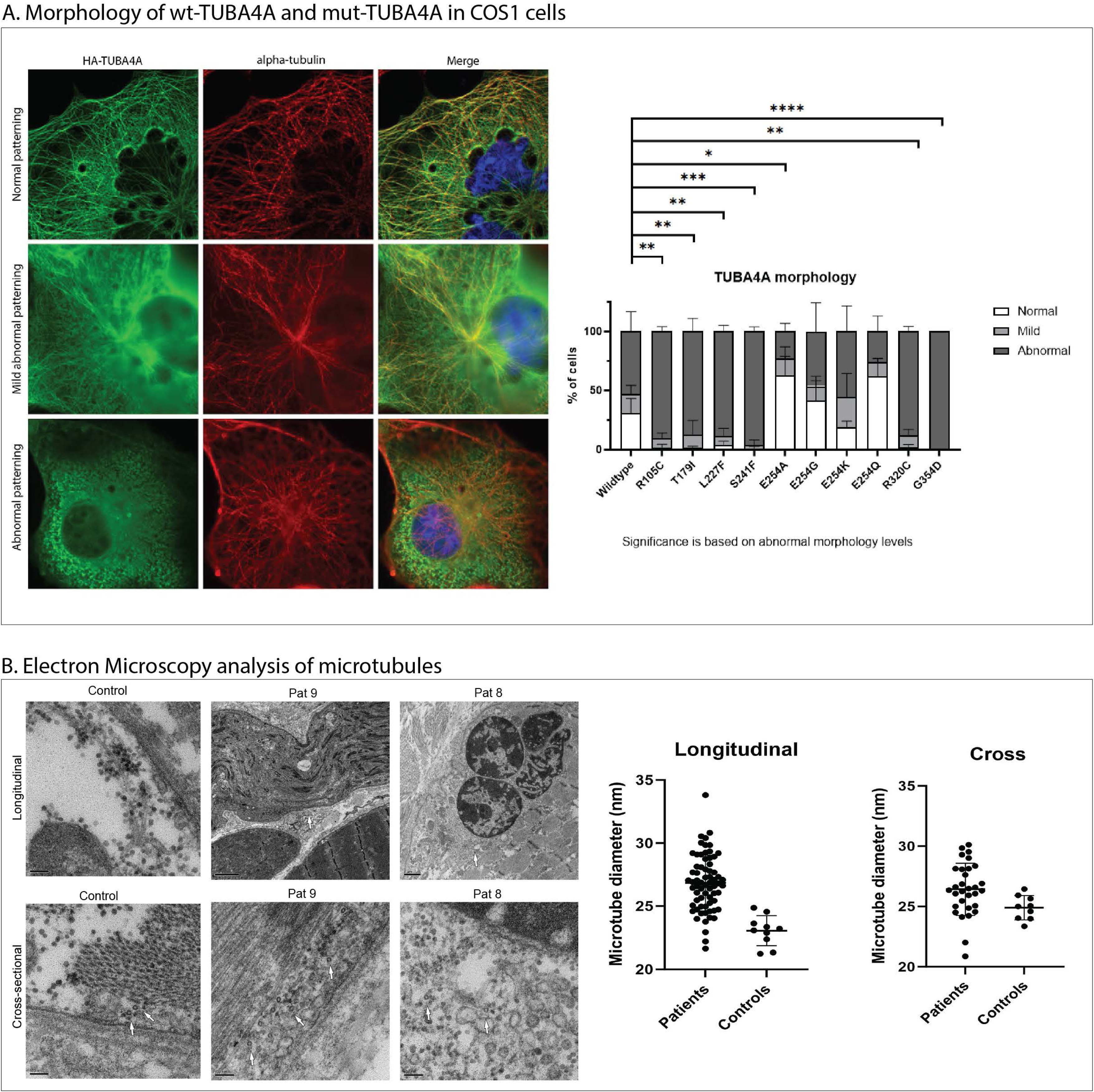
*TUBA4A* variants disrupt microtubule morphology in cellular models and patient muscles. A) Molecular physiology of cytoskeleton in COS1 cells overexpressed with TUBA4A-wildtype (wt) and TUBA4A variants. Morphology ranged from normal, mild to abnormal cytoskeleton. As compared to TUBA4A-wt, previously reported disease-causing variants Arg105Cys (ALS) and Arg320Cys (ALS) showed abnormal cytoskeleton structure. Additionally, the myopathic variants identified in our study, Thr179Ile, Leu227Phe, Ser241Phe and Gly354Asp showed abnormal cytoskeletal patterning. Conversely, Glu254Ala, Glu254Gly, Glu254Lys and Gln254Gln showed normal to mildly abnormal cytoskeletal patterning. B) Electron microscopy analysis of microtubules in control muscles and patient muscles showing statistically significant increase in microtubule diameter in patient muscles as compared to controls.

### Microtubule thickening in myo-tubulinopathies

To investigate potential ultrastructural differences in microtubules between patients and age matched controls, we performed additional electron microscopy investigations of patient muscle biopsies from Pat.8 (p.(Glu254Lys), 0-5-year-old male) and Pat.9 (p.(Glu254Gln), 0-5-month-old male) and compared them to two age-matched control biopsies (0-5-year-old males). This analysis revealed significantly thicker microtubules myofibres in patient muscle compared to age-matched healthy controls (*p* < 0.0001) (Fig 4B).

### Reassessment of identified *TUBA4A* variants

Based on the genetic data, *in silico* evidence, and findings from the *in vitro* assay it was possible to reclassify most of the variants in the myo-tubulinopathy cohort using the ACMG/AMP guidelines (Table 3, Supplementary Data 2).^40^ Accordingly, classification of eight VUSs could be upgraded to ‘pathogenic’ using additional ACMG/AMP criteria, while there was sufficient evidence to suggest upgrading three variants to ‘likely pathogenic. Two variants had insufficient evidence and remained as VUS (Table 3, Supplementary Data 2). Raw and revised classification and assertion criteria details can be seen in Supplementary Data 2.

## Discussion

In this multi-centre study, we identified 13 different missense variants in *TUBA4A* causing an axial myopathy, with childhood onset of disease being the most frequent clinical presentation. Consistent with our methods of ascertainment, which were largely focused on myopathic cases from neuromuscular disease cohorts, 17/19 families did not have a recorded history of motor neuron disorders, thus this represents an important phenotypic expansion of *TUBA4A*-related disease to primary skeletal muscle conditions. Two probands presented with multisystem involvement: a mild myofibrillar myopathy along with cerebellar ataxia, cerebellar atrophy and epileptic seizures. These two cases thus represent the first incidence of an MSP phenotype associated with pathogenic variants of *TUBA4A*. Both MSP associated variants occurred in exon 2 of *TUBA4A*, whereas the other 11 variants associated with pure myopathic phenotypes were clustered in the exon 4. Three probands harboured homozygous variants in *TUBA4A* consistent with the observed recessive phenotypes. Of these, two probands presented with severe, early-onset axial myopathy, while the third displayed a later-onset multisystem phenotype.

In general, tubulinopathies are either predominantly autosomal dominant (AD) or *de novo*/sporadic conditions and are rarely autosomal recessive.^41^ In our cohort, we observed four families with autosomal dominant inheritance; 12 cases had sporadic occurrence of a skeletal muscle phenotype where five probands had *de novo* variants in *TUBA4A*, while in nine isolated cases, we could not clarify the mode of inheritance due to absence of additional familial DNA. Of the remaining families, three showed clear autosomal recessive inheritance with affected individuals homozygous for bi-parentally inherited variants.

Notably, multiple substitutions affecting the same amino acid residue resulted in similarly severe myopathic phenotypes in our cohort. Four missense variants were observed at Glu254 (Glu254Gly, Glu254Lys, Glu254Ala and Glu254Gln) in four different probands, each time resulting in a severe early onset phenotype, including two who died in the first year of life. Thus, we observed a range of clinical features including different patterns of inheritance, varying severity of muscle weakness from death in the first year of life (n = 2), non-ambulation in childhood, to mild or moderate weakness in adulthood.

In patient muscles biopsies, histopathological findings included characteristic features of a myopathy such as variation in myofibre size, myofibrillar disorganisation, type 1 myofibre predominance, internal nucleation, regenerating and degenerating myofibres, autophagic vacuoles (*n* = 4), nemaline bodies (*n* = 4), minicores and moth-eaten myofibres. Ultrastructural studies showed classical signs of myofibrillar disarray (*n* = 4). Muscle biopsies from the two MSP cases showed similar myopathic features. Owing to the severity of the clinical presentation in affected individuals from the two recessive families, muscle biopsies could not be performed for diagnostic evaluation. In our cohort, however, there were no clear unifying histopathological features. Previously Wan and colleagues^15^ showed that in *TUBA4A*-p.(Leu227Phe) related myopathy, both probands exhibited myofibres with focal myofibrillar disorganisation and rimmed vacuoles.

Variable accumulation of phosphorylated TDP-43, often considered a marker of a proteinopathy, showed that different *TUBA4A* variants might result in distinct level of protein accumulations. Similarly, TUBA4A-positive accumulations were also increased in a subset of muscle biopsies compared to others and controls. Although none of the previous *TUBA4A*-related ataxia and ALS studies investigated specific autophagic disturbances in patient biopsies, Smith and colleagues^7^ showed accumulation of TUBA4A p.(Trp407Ter) in transfected primary motor neurons. Wan and colleagues^15^ showed ubiquitin-positive TUBA4A accumulations of the TUBA4A-p.(Leu227Phe) variant in transfected COS7 cells and suggested the involvement of autophagic disturbances. Interestingly, the same variant, p.(Leu227Phe), was observed in two cases within our cohort (*de novo* in Pat.7 and as an isolated sporadic occurrence in Pat.6). Complementing these earlier findings,^15^ we also observed classical signs of disturbed autophagy, i.e. p62/SQSTM1-positive accumulations in myofibres of affected patients. However, from our immunohistochemistry data and previous studies in tubulin-related pathomechanisms^7,42^, protein accumulations may not be the sole contributor to the pathobiology of the disease.

We therefore used a range of *in silico* methods to predict and analyse the molecular impact of identified variants in *TUBA4A*. Given that tubulin variants can disrupt either tubulin monomer dimerisation or microtubule stability through a dominant-negative mechanism, these computational approaches may offer insights into the distinct ways each variant might contribute to disease pathology.

3D modelling studies indicated that p.(Leu227Phe) showed considerable changes to polar contacts and steric hindrance upon substitution, which could affect protein stability. Further, our FoldX calculations also showed that p.(Leu227Phe) exhibited significantly elevated free energy changes (threshold 1.6 kcal/mol), indicating substantial protein destabilisation. Our *in vitro* studies in COS1 cells showed abnormal microtubule morphology in cells transfected with the p.(Leu227Phe) variant. These observations are in alignment with Wan and colleagues^15^ who showed that the TUBA4A:p.(Leu227Phe) variant forms ubiquitin-positive TUBA4A accumulations in transfected COS7 cells. Notably, while FoldX predicted severe destabilisation, our *in-silico* aggregation propensity analysis did not suggest an increased tendency toward aggregation for this variant compared to wild-type TUBA4A. This apparent divergence illustrates how distinct molecular mechanisms can lead to similar cellular phenotypes. Our integrated analysis suggests that the p.(Leu227Phe) substitution introduces a bulkier aromatic side chain in a spatially constrained region of TUBA4A, creating specific steric clashes with its neighbouring residues. These structural perturbations directly disrupt the tightly packed hydrophobic interactions that stabilise this domain, accounting for the observed significant increase in free energy. Thus, p.(Leu227Phe) contributes to pathology likely through the thermodynamic destabilisation of the protein’s native fold which may impair proper degradation of the protein resulting in improper localisation and accumulation of the mutant protein rather than through increased aggregation propensity.

The p.(Glu254Ala) and the p.(Glu254Gly) variant displayed increased aggregation propensity compared to the wild-type TUBA4A, while our FoldX calculations showed that all identified variants at this hotspot [p.(Glu254Ala), p.(Glu254Lys), p.(Glu254Gln), and p.(Glu254Gly)] have relatively minimal changes in free energy. Interestingly, *in vitro* over-expression studies revealed normal to mild morphological changes in cells expressing these four variants. This contrasts with the clinical presentation, as patients harbouring variants at this position exhibited the most severe phenotypes in our cohort. Notably, muscle biopsies of Pat.8 with p.(Glu254Lys), Pat.9 with p.(Glu254Gln) and Pat.11 with p.(Glu254Ala) showed TUBA4A positive protein accumulations (Fig 2B:x-xii). Pat.8 with p.(Glu254Lys), Pat.9 with p.(Glu254Gln) also showed significantly dilated microtubules in skeletal muscle compared to the controls (Fig 4B). Again, this divergence between computational predictions, cellular phenotypes, and clinical severity highlights the complex nature of these variants and the limitations of *in vitro* over-expression assays. While variants at this hotspot may promote aggregation, and possibly impact tubulin dimerisation, the exact pathomechanism underlying the noteworthy clinical impact of this mutational hotspot requires further investigation. Recently, Dodd and colleagues observed similar heterogeneity in tubulin pathologies associated with specific *TUBB4B* variants, across *in silico*, *in vitro* and *in vivo* studies.^42^

The p.(Glu284Lys) variant identified in five unrelated families and 13 affected individuals with similar mild myopathic presentation in our myo-tubulinopathy cohort, presents a notable case of phenotypic divergence. This variant has previously been reported in relation to human zygotic arrest and early developmental failure in heterozygosity.^14,43^ All cases in our cohort, harbouring this heterozygous variant, either showed autosomal dominant inheritance (Pat.15, Pat.16A, Pat.16B, Pat.17, and Pat.18) or had an undetermined inheritance (Pat.13 and Pat.14), and none of the affected females reported infertility or reproductive complications. Conversely, none of the individuals in the fertility studies^14,43^ were reported to have neuromuscular disease (personal communication), suggesting potential tissue-specific effects of the variant. However, this pleiotropic effect is not unique to *TUBA4A*, as similar divergent phenotypes have been observed with other variants such as *TUBB3*:p.Arg262His - implicated in cortical malformations, peripheral neuropathy and endocrine malfunction,^44^ and *LMNA*:p.Arg644Cys, which has been associated with muscular dystrophy, cardiomyopathy, insulin resistance, lipodystrophy and peripheral neuropathy.^45^

In Pat.21 (p.(Gly354Asp)) and the siblings Pat.22 and Pat.23 (p.(Gly354Asp)) we observed an autosomal recessive myo-tubulinopathy, due to homozygous pathogenic variants. Interestingly, these three patients also presented with fatigability. Repeat nerve stimulation (RNS) showed decrements after repetitive stimulation of ulnar nerve and accessory nerve (Supplementary Data 1(available upon request)). The clinical presentation of fatiguability and decrement on RNS prompted treatment with pyridostigmine. Both patients showed improved muscle function post-treatment. Pat.7 with the *de novo* p.(Leu227Phe) variant and Pat.25 with the *de novo* p.(Gly350Val) variant also showed similar fatigability. Pat.7 showed decrement of up to 17% in ulnar nerve and was prescribed salbutamol 0.1 – 0.3 mg/kg/day for 2-3 months but showed no improvement. For Pat.25 there was no decrement observed on RNS of ulnar and ocular muscle but from deltoideus (up to 22%). However, she did not respond to the salbutamol treatment as well. This aligns well with our integrated findings that specific molecular consequences unique to each *TUBA4A* variant and their distinct downstream interactions may exist in different myo-tubulinopathies, ultimately determining the potential therapeutic approaches and outcomes.

In our cohort, 17/19 families did not show a history of a motor neuron disease, and the phenotype observed was primary skeletal muscle disease. However, in these patients, especially with earlier onset and the young ages within the cohort, involvement of CNS cannot be excluded at a later age.

Interestingly, in Pat.24 harbouring the heterozygous likely-pathogenic, p.(Gln11His), we observed a mild early-onset cerebellar atrophy along with a myofibrillar myopathy (MFM). In Pat.26 harbouring the homozygous VUS, p.(Ala12Thr), pathologically confirmed MFM and concomitant late-onset ataxia with cerebellar atrophy was observed.

The positively charged Gln11 residue interacts with the α-tubulin catalytic GTP site. A change to the negatively charged His at this position will potentially inhibit the GTP binding activity and hence could impact microtubule dynamics. In the α/β-tubulin heterodimer, α-tubulin has a non-hydrolysing and non-exchangeable “N-site” which binds to GTP, whereas β-tubulin has an exchangeable “E-site” where GTP can be hydrolysed into GDP.^46,47^ Recently, McFadden and colleagues reported the p.(Glu11Arg) variant in *TUBB4B*, observed in a syndromic presentation of hearing loss, hyperopia without retinal abnormalities, renal tubular Fanconi Syndrome and hypophosphatemic rickets. Functional studies showed that this p.(Glu11Arg) variant results in inhibition of GTPase activity due to the Arg change that impedes depolymerisation and hence results in hyperstability of the polymerised microtubule.^48^ The hydrophobic Ala12 residue is close to the α-tubulin GTP binding site, and similarly a change to a polar uncharged Thr residue at this position might impact the GTP binding activity of the heterodimer. Notably, the Gln11His variant was identified as a heterozygous *de novo* change, whereas the Ala12Thr variant was homozygous. Despite these distinct zygosities, both variants are associated with similar MSP phenotypes, raising intriguing questions about the mechanistic convergence of their allelic presentations.

Pat.26 with the homozygous VUS p.(Ala12Thr) was previously published with a VUS in *FLNC* thought to be the cause of the myofibrillar myopathy in the proband.^49^ After the publication, the *FLNC* variant (p.(Thr2419Met)) has been reported as ‘likely benign’ in multiple submissions in ClinVar (VCV000712192.18). Therefore, with the current knowledge it is unlikely to be the (unique) cause of the disease in the proband, although a potential role in modulating the phenotype cannot be excluded. From our results, we believe that these two substitutions, *TUBA4A* p.(Gln11His) and p.(Ala12Thr), are quite distinctive in terms of their clinical presentations involving multiple systems, possibly due to the unique biochemical changes to the GTP-binding site in TUBA4A.

In general, MSPs encompass a group of inherited disorders characterised by the degeneration of muscle, bone, and nervous tissues.^50^ A disease presentation can be classified as an MSP if a neurodegenerative and a neuromuscular phenotype with protein aggregations coexist in the same individual or co-segregate with a variant in the same family.^50,51^ The shared pathology involves disruptions in key protein clearance pathways, specifically the ubiquitin-proteasome system (UPS) and autophagy^50^ - leading to accumulation of toxic protein aggregates, manifesting as diverse clinical phenotypes such as rimmed vacuolar myopathy (RVM), Paget disease of bone (PDB), FTD, and ALS.^50^ Essential components of the two major protein quality control pathways: UPS and autophagy, including sequestosome 1/p62 (SQSTM1),^52^ play pivotal roles in MSPs.^50^

*TUBA4A* variants have been implicated in neurodegenerative diseases such as ALS,^7,53–55^ FTD,^8,9,56^ and ataxia.^11^ In our study, p62 positive accumulations are observed in all patients in whom p62 staining was performed (n = 5). Wan and colleagues also showed presence of p62 positive TUBA4A accumulations in COS-7 cells.^15^ The observation of an MSP phenotype in Pat.24 (p.(Gln11His)) and Pat.26 (p.(Ala12Thr)) in our cohort suggests that *TUBA4A*-related MSPs may represent a unique subtype, warranting further investigation into clinical and molecular features associated with TUBA4A variants observed in MSPs.

The diverse neurodegenerative and neuromuscular phenotypes associated with TUBA4A - and tubulins more broadly - can be partly explained by the ‘tubulin code’. This concept proposes that the functional diversity of microtubules, across tissues, is regulated by the expression of distinct tubulin isotypes and a range of post-translational modifications (PTMs), including acetylation, polyglutamylation, and detyrosination.^57,58^ These PTMs fine-tune microtubule dynamics, stability, and interactions with microtubule-associated proteins, tailoring microtubule behaviour to specific cellular contexts.^59^ In skeletal muscle, where microtubules contribute to mechanical stability, nuclear positioning, and metabolic regulation, microtubule regulation is especially critical.^60^ Disruption of TUBA4A in this context may therefore have a more pronounced effect on microtubule function, directly impairing myofibre integrity and contractility.

Recently, Dodd et al.^42^ experimentally showed how the tubulin code explains phenotypic diversity associated with *TUBB4B* variants. Based on this increasing evidence and our findings, we propose that the tubulin code plays a fundamental role in determining the spectrum of neurodegenerative and neuromuscular manifestations observed in *TUBA4A*-related disorders, offering a plausible mechanism for the diverse clinical presentations observed.

The publicly available GTEx^61^ and Human Protein Atlas^62^ datasets show ubiquitous expression of alpha-tubulin 4a across human tissues, with notably high expression in skeletal muscle.

The wide range of disease onset, variable clinical manifestations and histopathologies, and multiple inheritance patterns observed in the cohort make diagnosis of myo-tubulinopathies and multisystem proteinopathies challenging. Importantly, all disease-causing variants identified in this study were missense changes, many of which would be classified as VUSs under current clinical guidelines owing to limited functional evidence and the absence of an established, reproducible gene-disease relationship for *TUBA4A* in skeletal muscle disorders. This highlights a critical gap: pathogenic *TUBA4A* variants may remain unreported or misclassified, delaying diagnosis and hindering appropriate clinical follow-up.

In conclusion, this study establishes that missense variants in *TUBA4A* can cause myopathy and multisystem proteinopathy with variable clinical and histopathological features, including p62 accumulation, internal nucleation, autophagy dysfunction, and myofibrillar disarray. These findings support a role for *TUBA4A* in maintaining skeletal muscle integrity through the regulation of microtubule function and dynamics. Our large, multi-centre cohort expands the phenotypic and inheritance spectrum of *TUBA4A*-related disease, with most patients showing no evidence of CNS involvement. Together, these results underscore the importance of including *TUBA4A* in genetic screening panels for unresolved myopathies and related neuromuscular conditions.

## Data Availability

All relevant imaging and biological materials included in this study are available from the authors (or commercial providers). Sensitive sr-ES/sr-GS data of patients and family members are deposited in seqr and RD-Connect GPAP.

## Acknowledgements

The authors would like to thank the patients and their families for cooperation in this study. Wathone Win and Ronald van Beek for technical assistance. Clinical and genetic data of the most patients were shared in RD-Connect, which received funding from the European Union Seventh Framework Programme (FP7/2007-2013) under grant agreement No. 305444 within the Solve-RD project. Solve-RD project has received funding from the European Union’s Horizon 2020 research and innovation programme under grant agreement No 779257. This work was supported by resources provided by the Pawsey Supercomputing Research Centre with funding from the Australian Government and the Government of Western Australia.

## Author contributions

Conceptualisation of the study: MJ, GR

Project administration: MJ, GR

Funding acquisition: MJ, GR, NGL

Patient samples and clinical/genomic data collection: All clinicians and molecular geneticists Data analysis and curation: MJ, CF, JMP, MMO, YS, MA, EYZ, HRL, CCB, SAS, EAE, LMK, PBK, IOO, BM, LP, MOL, CAT, AODL, FCL, ADA, AC, MT, AP, CVG, GS, MLMN, MC, CAG, ZMV, AHB, VM, GK, AK, AT, NSP, JV, CGC, EZ, CMM, MS, GT, VS, BU, IN, GR

Methodology: MJ, YS, MMO, JMP, GR

Visualisation: MJ, YS, MMO, JMP, KSS, IAC, DA, SK, CMM, GS, NSP, JV

Writing the original draft: MJ, YS, MMO, JMP, GR

Review and editing of the manuscript: All authors

---

**Supplementary Table.**
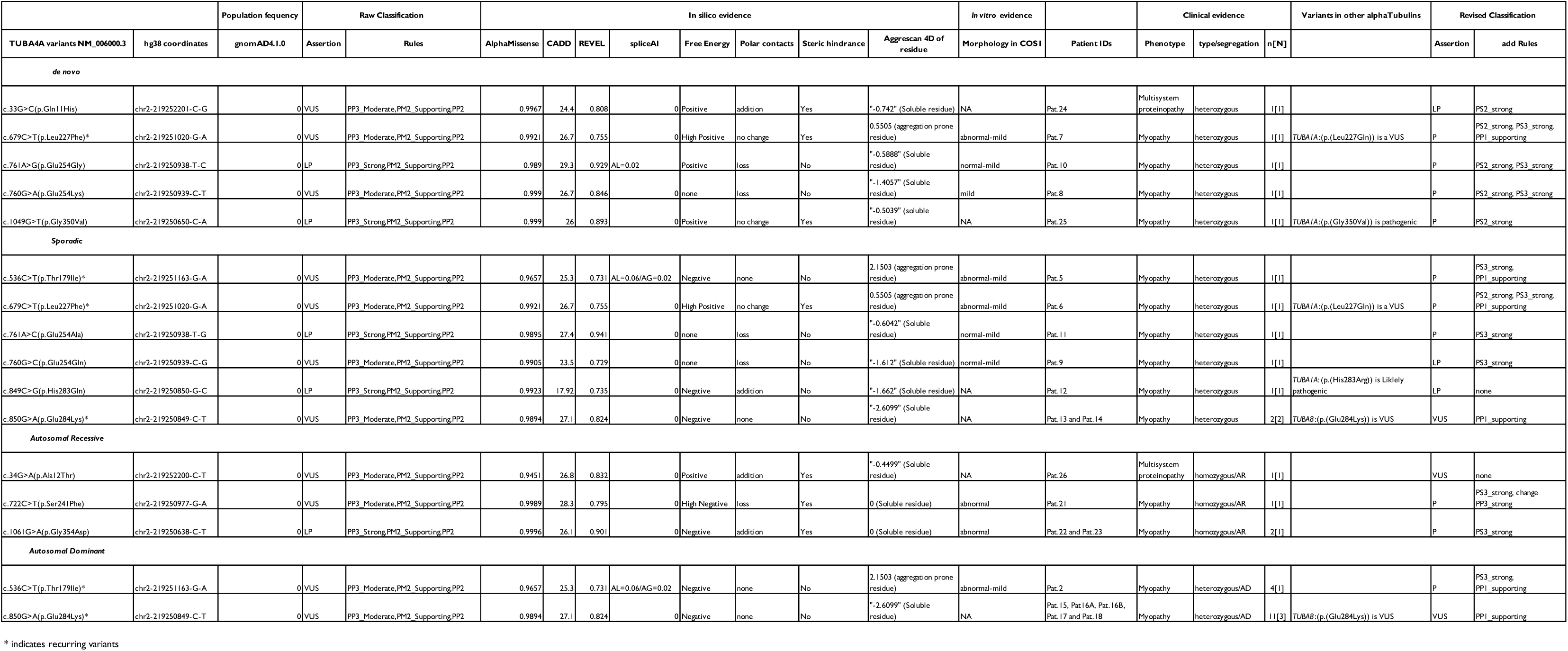
Assessment of variants in TUBA4A observed in our cohort. AL=Acceptor Loss, AG=Acceptor Gain, AR = Autosomal recessive, AD = Autosomal dominant, n=number of confirmed affected individuals, N=number of families, NA=Not assessed.

